# Impact of a Clinical Decision Support Intervention on Albumin Utilization and Appropriateness of Use in an Academic Healthcare System

**DOI:** 10.1101/2021.04.05.21254943

**Authors:** Prashant R. Mudireddy, Nikhil K. Mull, Kendal Williams, Jennifer Lukaszewicz Bushen, Nishaminy Kasbekar, Karen Krok, Asaf Hanish, Benjamin French, Craig A. Umscheid

## Abstract

**Background:** Albumin is expensive compared to crystalloid intravenous fluids and may be used for inappropriate indications, resulting in low value care.

**Aim/Purpose:** To study the impact of a computerized clinical decision support (CDS) intervention on albumin utilization and appropriateness of use in an academic healthcare system.

**Methods:** A systematic review examining appropriate indications for albumin use in the healthcare setting was used by an interprofessional group of stakeholders locally to develop a CDS intervention to improve the appropriateness of albumin utilization. The order set was implemented across our healthcare system on 4/12/2011, included a list of appropriate indications, and automatically provided albumin concentration, dose and frequency based on the indication selected and patient weight and creatinine. We measured units of albumin ordered across the healthcare system and individually at each of three hospitals in the healthcare system 12 months before and after intervention implementation. An interrupted time series analysis using monthly data examined changes in the level and slope of albumin use during pre-versus post-implementation periods. We also reviewed charts of all adult inpatients receiving albumin in the 3 months prior to and following implementation of the order set at two of the three hospitals within the healthcare system, to determine if “appropriateness” of use had changed, as defined by our consensus criteria. We selected the two hospitals with the most frequent use of albumin in the pre-period. We used chi square tests to compare changes in the proportion of appropriate instances and grams of albumin used. We considered a p-value <0.05 as statistically significant.

**Results:** The number of patient encounters analyzed in the 12 months before and after the albumin CDS intervention was 79,108, and 78,240, respectively. There was a statistically significant decrease in mean units of albumin ordered immediately post-intervention across the healthcare system (−4.98 units per 1000 patient days, confidence interval −9.64 to −0.33, p=0.04). At Hospital 1, there were no statistically significant changes in albumin ordering over time. At Hospital 2, albumin ordering significantly increased up to the intervention, but decreased significantly immediately following the intervention and continued to decrease significantly over time following the intervention; the pre and post implementation slopes were significantly different. At Hospital 3, albumin ordering was statistically unchanged up to the intervention, decreased significantly immediately following the intervention, and significantly increased over time following the intervention, but the pre and post slopes were not statistically different. At Hospitals 1 and 3, there was a statistically significant improvement in “appropriateness” of albumin use in the three months following implementation.

**Conclusions:** Implementation of a CDS intervention was associated with an increase in the amount of albumin administered appropriately at two hospitals within an academic healthcare system and an overall decrease in albumin utilization across the healthcare system.

## Introduction

Human serum albumin is a multifunctional, non-glycosylated, negatively charged protein that accounts for over 50% of total plasma protein (1). It is responsible for maintaining colloid osmotic pressure. In addition, it has other properties including ligand and metal binding, reactive oxygen and nitrogen species scavenging, and anti-inflammatory effects (1). The appropriate use of albumin in hospitalized patients is frequently debated. A meta-analysis published by Cochrane in 1998 (2) suggested that mortality increased in patients receiving albumin. However, a subsequent meta-analysis published by Wilkes and colleagues in 2001(3) contradicted this and supported the safety of albumin. In another meta-analysis by Vincent and colleagues in 2004 (4), albumin was shown to reduce morbidity in acutely ill hospitalized patients. In that same year, the “Saline versus Albumin Fluid Evaluation (SAFE)” trial suggested that in critically ill patients, normal saline was as effective as albumin when used as a resuscitation fluid with no differences between groups in morbidity, length of intensive care unit (ICU) or hospital stay, and survival (5).

The use of albumin is associated with risks including anaphylactic reactions (6,7) as well as theoretical risks of transmission of infections (including prions). In addition, albumin is expensive relative to crystalloid intravenous fluids. Therefore, it is important that albumin be used appropriately to ensure safe and high value care.

The indications for and use of albumin have undergone substantial changes in the past several years. For example, previously, administration of albumin was considered appropriate for the management of shock with hypoalbuminemia as well as for paracentesis >1.5 L (8,9); however, recently published guidelines have considered these indications as inappropriate (10,11). These changes have likely contributed to poor compliance with established or entity specific guidelines and increasing proportions of inappropriate albumin use across both the US and Europe (12,15-22,24,25).

We hypothesized that the development and implementation of a computerized clinical decision support (CDS) intervention targeting albumin use would reduce overall and inappropriate albumin use. The primary aim of this study was to assess the utilization and appropriateness of albumin use in an academic healthcare system following the implementation of such an evidence-based order set.

## Methods

We developed an electronic order set to guide albumin ordering. We subsequently evaluated the impact of this CDS intervention on albumin use at the 3 hospitals within our academic healthcare system, one of which (Hospital 1) is a liver transplant center.

### Development of CDS intervention

A systematic review of the literature was used to summarize the evidence for appropriate indications for albumin use in the healthcare setting. The review examined electronic bibliographic databases such as MEDLINE, EMBASE, and Cochrane, as well as websites of relevant societies including the American Thoracic Society (ATS), American Association for Study of Liver Diseases (AASLD), Society of Critical Care Medicine, American Association of Nephrology, and the American College of Surgeons. The review informed a list of appropriate indications for albumin use, which was then vetted by clinician stakeholders representing relevant clinical departments as well as pharmacy. An electronic order set based on this consensus list was designed and implemented in the electronic health record (EHR) (Sunrise Clinical Manager 5.5, Allscripts, Chicago, IL) used across the inpatient services of our multi-hospital academic healthcare system (Figure 1).

**Figure 1.**
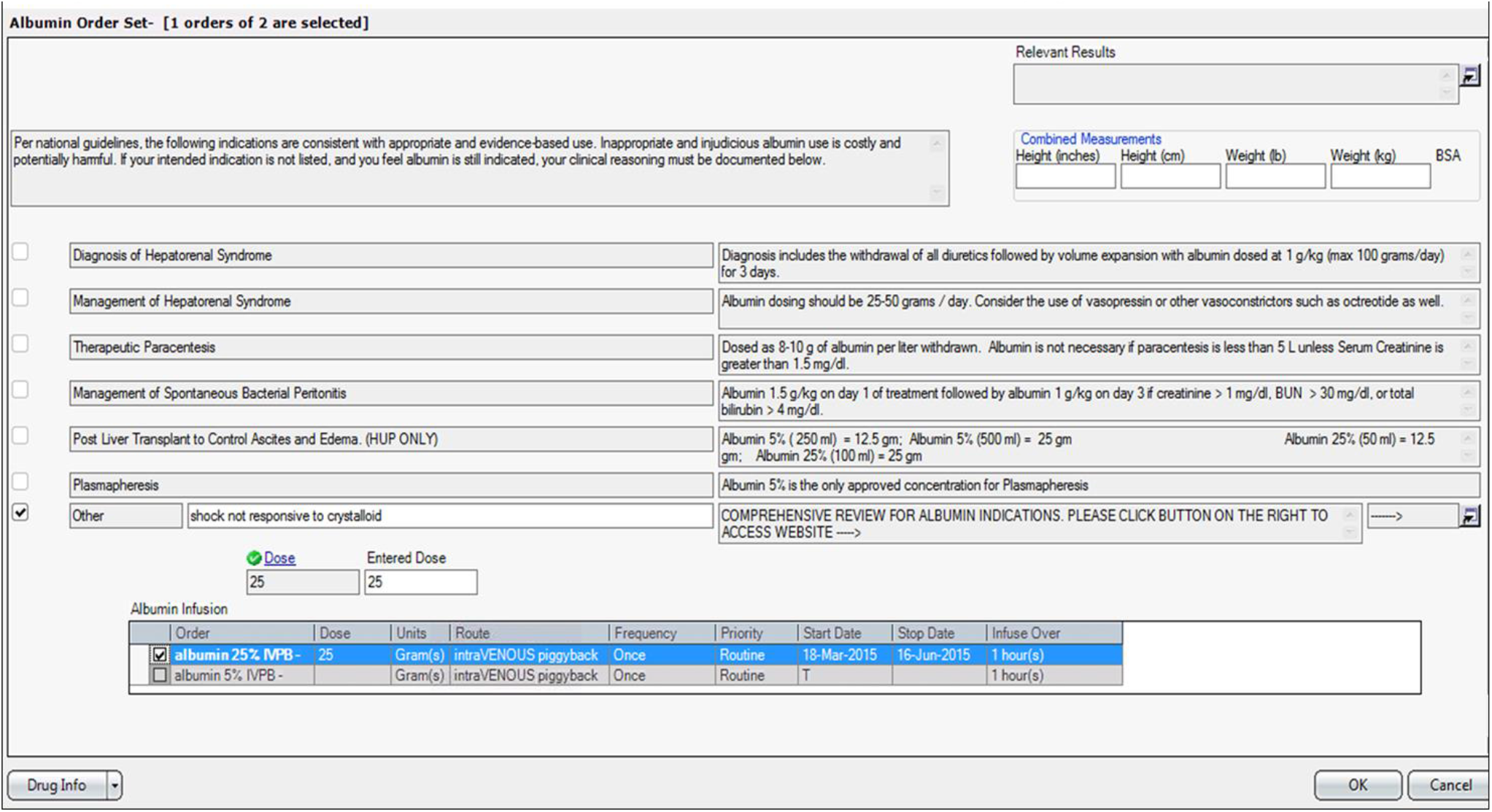
Albumin clinical decision support order set. The goal of the order set was to guide practitioners in the appropriate use of albumin by displaying appropriate indications for use, and linking these indications to the appropriate concentrations, doses, and frequencies of albumin. An automated dose calculator prevented patients from getting incorrect doses. A link within the order set enabled access to an evidence summary. To avoid “hard stops” and allow for clinician judgement, the indication category also allowed providers to select “other” as needed.

Prior to the implementation of the albumin order set, albumin could be ordered through the alpha browse of our EHR. The Department of Pharmacy and the Pharmacy and Therapeutics Committee had outlined appropriate use guidelines which were available on a separate website. These guidelines were intended to guide therapy. Enforcement of guidelines was dependent on individual pharmacy practitioners.

The intent of the order set was to guide practitioners in the appropriate use of albumin by displaying appropriate indications for use, and linking these indications to the appropriate concentrations, doses, and frequencies of albumin. When the electronic order set was implemented (April 12, 2011), the alpha browse option was removed so that providers were only able to order albumin through the order set. The order set also limited certain concentrations of albumin to specific indications based on the systematic review. An automated dose calculator was included in the order set to prevent patients from getting incorrect doses. Due to the relative expense of the product and how it is supplied from the manufacturer, rounding to the nearest bottle size was also implemented to prevent waste. A link within the order set was available to access a summary of the evidence so that providers did not need to go outside of the order set to view the evidence review or guidelines. To avoid “hard stops” and allow for clinician judgement, the indication category also allowed providers to select “other” if they needed the agent for an indication other than one listed.

### Evaluation of the impact of the CDS intervention

To study the impact of the order set on albumin utilization, we examined units of albumin ordered in the 12 months before and after CDS implementation (April 1, 2010 to March 31, 2012). We then performed a chart review of all inpatients receiving albumin at two of the hospitals within the healthcare system in the 3 months before (January 1, 2011 to March 31, 2011) and after (May 1, 2011 to July 31, 2011) order set implementation to examine changes in the “appropriateness” of albumin use, as defined by our consensus criteria. For our analysis of “appropriateness”, we selected the two hospitals with the most frequent use of albumin in the pre-period. The study received expedited approval and a Health Insurance Portability and Accountability Act (HIPAA) waiver from the University of Pennsylvania Institutional Review Board prior to data collection.

### Data collection

Our EHR and financial database (Horizon Performance Manager) provided data for the analysis. The relevant inpatient encounter for all adult inpatients included in our “appropriateness” chart review was examined by a single reviewer for each of the two hospital sites. Data collected included patient demographics, indications for albumin use, appropriateness of albumin use, ordering department, doses of albumin ordered, and albumin concentration. “Appropriate use” was defined as use per an indication in the order set which was supported by documentation in the chart. “Inappropriate use” was defined as albumin use for an indication not listed in the electronic order set, or use per chart documentation that is not congruent with the indication selected in the order set. “Indeterminate” was defined as the inability to establish a reason for albumin use based on chart review.

Each time a patient received albumin for a particular indication, it was counted as an “order”. Thus, a patient receiving albumin for two different indications (for example, for the treatment of hepatorenal syndrome [HRS] and post-paracentesis) would be counted as two separate orders.

### Statistical Analysis

We examined units of albumin ordered per 1000 patient days in the 12 months before and after CDS implementation using interrupted time series analyses for the healthcare system as a whole, as well as the individual hospitals. Analyses examined changes in the level and slope of albumin use during pre-versus post-implementation periods, using monthly data. We used chi square tests to compare changes in the proportion of appropriate instances and grams of albumin used. We considered a p-value <0.05 as statistically significant. Analyses were performed using Stata software (Stata Corp, College Station, TX).

## Results

The number of patient encounters analyzed in the 12 months before and after the intervention was 79,108, and 78,240, respectively. There was a statistically significant decrease in mean units of albumin ordered immediately post-intervention across the healthcare system (−4.98 units per 1000 patient days, confidence interval −9.64 to −0.33, p=0.04). (Table 1, Figure 2) At Hospital 1, there were no statistically significant changes in albumin ordering. At Hospital 2, albumin ordering significantly increased up to the intervention and decreased significantly immediately following the intervention; the pre and post implementation slopes were significantly different. At Hospital 3, albumin ordering was statistically unchanged up to the intervention and decreased significantly immediately following the intervention, but the pre and post slopes were not statistically different. (Table 1, Figure 2)

**Table 1.** **Change in albumin ordering pre and post order set implementation. Slopes correspond to average per-month change in albumin units ordered per 1000 patient-days. Immediate effect corresponds to difference in albumin units ordered per 1000 patient-days immediately after the intervention.**

| **Hospital** | **Slope Before Intervention: $\beta$ (95% CI), p** | **Immediate Effect: $\beta$ (95% CI), p** | **Slope After Implementation: $\beta$ (95% CI), p** | **Difference Between Slopes** | **Pre-Post Means\*** |
| --- | --- | --- | --- | --- | --- |
| **1** | -0.14 (-0.88 – 0.60), 0.69 | -0.21 (-7.46 – 7.05), 0.95 | 0.96 (-0.08 – 2.01), 0.07 | Not Significant | +4.35 |
| **2** | 0.92 (0.28 – 1.55), <0.01 | -7.60 (-13.79 - -1.39), 0.02 | -0.97 (-1.86 - -0.07), 0.04 | Significant | -2.96 |
| **3** | -0.55 (-1.47 - 0.37), 0.23 | -16.34 (-25.38 - -7.30), <0.01 | 1.53 (0.23 – 2.83), 0.02 | Not Significant | -13.12 |
| **All** | 0.06 (-0.41 – 0.54), 0.14 | -4.98 (-9.64 - -0.33), 0.04 | 0.55 (-0.12 – 1.22), 0.10 | Not Significant | -0.61 |

CI: confidence interval.
\*Pre-Post Difference between Mean Units of Albumin Ordered per 1000 Patient Days

**Figure 2.**
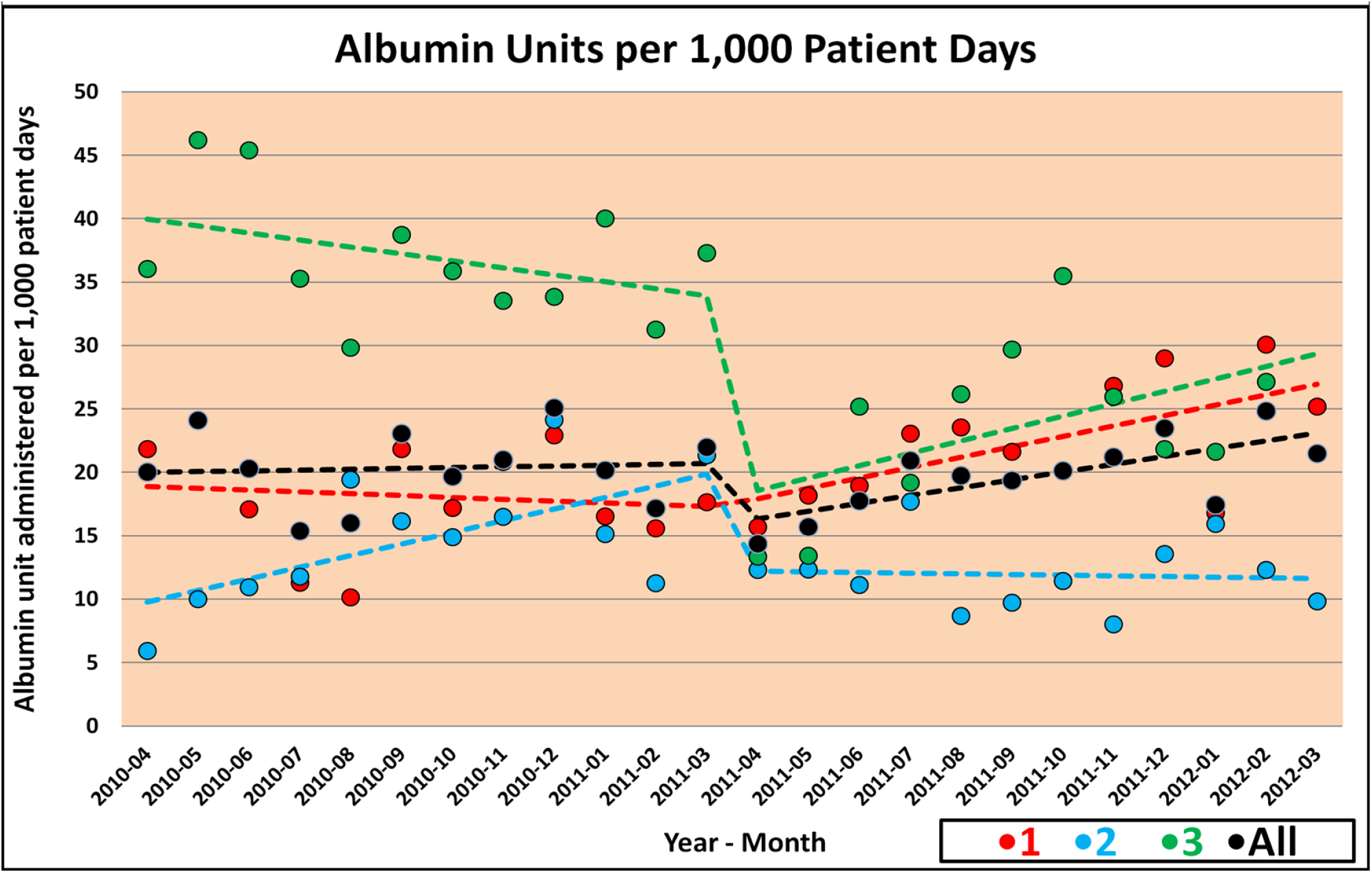
**Interrupted time series analysis of albumin ordering 12 months pre and post albumin order set implementation**

The differences in appropriateness of albumin orders and grams administered in the three months before and after the intervention at Hospitals 1 and 3 are described in Table 2.

**Table 2.** **Appropriateness of albumin administration 3 months pre and post albumin order set implementation at Hospitals 1 and 3**

|  | **Pre** | **Post** | **P value** |
| --- | --- | --- | --- |
| **Hospital 1** |  |  |  |
| Instances of albumin use (n) | 205 | 293 | - |
| Total appropriate albumin instances (n) | 98 | 158 | - |
| Proportion of appropriate albumin orders | 48% | 54% | NS |
| Total albumin given (g) | 19,603 | 25,413 | - |
| Total appropriate albumin (g) | 9,213 | 18,806 | - |
| Proportion of appropriate grams | 47% | 74% | <0.001 |
| **Hospital 3** |  |  |  |
| Instances of albumin use (n) | 237 | 133 | - |
| Total appropriate albumin instances (n) | 172 | 113 | - |
| Proportion of appropriate albumin orders | 72% | 85% | <0.01 |
| Total albumin given (g) | 10,998 | 5,400 | - |
| Total appropriate albumin (g) | 8,473 | 4,850 | - |
| Proportion of appropriate grams | 77% | 90% | <0.01 |

There were increases in the appropriateness of both albumin orders and grams administered at both hospitals in the three months following the intervention, although the increase in appropriate orders at Hospital 1 was not statistically significant.

At Hospital 1, gastroenterology (40.5% and 31.7% pre and post order set, respectively) and transplant surgery (27.8% and 32.4% pre and post order set, respectively) ordered albumin most frequently. At Hospital 3, cardiothoracic surgery (66.7% and 84.2% pre and post order set, respectively) and thoracic surgery (11.0% and 5.3% pre and post order set, respectively) ordered albumin most frequently.

Management of the complications of liver cirrhosis (prevention of post-paracentesis circulatory dysfunction, and management of HRS and spontaneous bacterial peritonitis [SBP]) were the most common indications for albumin orders at Hospital 1. They accounted for 51.7% of orders pre-order set and 63.1% of orders post-order set implementation. The treatment of shock was the most common indication for albumin use at Hospital 3 (76.6% and 89.8% pre and post order set implementation, respectively).

## Discussion

Our study found that after introduction of an albumin order set, there was a reduction in albumin units ordered across our healthcare system, as well as an increase in the appropriateness of use. Although the proportion of grams “appropriately” ordered statistically improved at both hospitals examined, the improvement in the proportion of “appropriate” orders was only statistically significant at one of the two hospitals examined. The reason for this could be that the CDS intervention not only provided information about the appropriate indications, but also helped calculate the appropriate dosage based on indication selected.

We found that management of the complications of liver cirrhosis (e.g., prevention of post-paracentesis circulatory dysfunction and management of HRS and SBP) were the most common indications for albumin use at our liver transplant center (Hospital 1). In this same hospital, the number of “appropriate” albumin orders and grams administered increased following implementation. This is not surprising given that many randomized controlled trials evaluating the appropriateness of albumin use in the last two decades have been performed in liver disease (28-34), and many professional societies (including the AASLD, European Association for Liver diseases [EASL], and the International Ascites Club [IAC]) have issued guidelines for the appropriate use of albumin in liver disease (11,35,36).

Treatment of shock was the most common indication for albumin use at Hospital 3, which performs a large number of cardiothoracic surgeries in our healthcare system. This same hospital demonstrated a reduction in the number of albumin orders and grams administered following the intervention implementation, along with an increase in “appropriateness” of use. This is not surprising given that large randomized controlled trials have shown in the last decade that saline can be equally effective as albumin when used as a resuscitation fluid, making the use of albumin for shock frequently inappropriate (5). In fact, guidelines by various professional societies have recommended the use of saline for resuscitation over albumin, with albumin being recommended only when patients have received maximal doses of crystalloids, or there is contraindication for use of crystalloids (37,38).

In a study similar to ours at an academic hospital in Italy, Mirici-Cappa and colleagues (27) assessed the impact of guidelines on albumin utilization using an order form listing the recommended indications. They noticed a 15-20% decrease in albumin consumption following implementation of the guideline. The reduction remained stable over six years of follow-up. Similar to our study results, complications of cirrhosis were common indications for albumin use. But in their study, refractory ascites was the most common indication for albumin use in patients with cirrhosis (56.4%). Using albumin to treat refractory ascites is controversial; though it is approved in Italy, it is not endorsed by either AASLD or EASL (11,35). Our study considered albumin use for refractory ascites as inappropriate and few patients received it for this indication. Though the inappropriate use of albumin in their study was < 10%, this would have been greater if refractory ascites had been considered inappropriate. Also, this study assumed that data recorded on the order form accurately described the indication for albumin use; this was not confirmed by chart review as in our study.

Despite the focus on appropriate utilization of albumin by various national and international professional societies through guideline publication, the number of inappropriate albumin orders and amount of albumin (in grams) used inappropriately was fairly high at our institution. Prior to order set implementation, 52% and 28% of orders were inappropriate at Hospitals 1 and 3, respectively. Similarly, 53% and 23% of grams administered were inappropriate pre-order set at Hospitals 1 and 3, respectively.

Although we identified significant inappropriate albumin use in our study, compared to the two major U.S studies published on albumin utilization, our study had a greater percentage of appropriate albumin use both pre- and post-order set implementation, despite the fact that our guidelines were comparatively more stringent. In 1995, Yim and colleagues in a study of 15 U.S. academic medical centers found that most of the albumin use was inappropriate when evaluated against model guidelines (21). Similarly, a 2003 study by Tanzi and colleagues evaluating the appropriateness of albumin use in adult and pediatric patients (in both community and academic hospitals in the U.S.) found that only 28.2% of albumin was prescribed appropriately (25). Both studies used University Health Consortium (UHC) guidelines (10) to evaluate appropriateness of albumin use.

Although previous studies have demonstrated improved albumin utilization with educational interventions targeting physicians (13,14,17,23), our CDS intervention was unique because it included the appropriate indications within a computerized order set with a link to the evidence-based literature review used to develop the order set, in an effort to provide “just-in-time” education to physicians using the order set. In addition, incorporation of guideline recommendations into order sets has been shown to increase guideline compliance (41). Although the number of appropriate orders and grams administered did increase post-order set implementation, there remained inappropriate use following order set implementation. This suggests a potential role for more formal educational programs for both attending physicians, housestaff and advanced practice providers, directed towards those departments where the most inappropriate use occurs.

Our study had limitations. First, this was a retrospective study. Second, we assessed the effect of the order set immediately after its introduction across the healthcare system, so its long-term effects are unknown. Third, we did not study the impact of these guidelines on clinical outcomes (i.e., the effect on morbidity and mortality of patients).

In conclusion, implementation of a CDS intervention was associated with an increase in the amount of albumin (in grams) administered appropriately at two hospitals within an academic healthcare system and an overall decrease in albumin utilization across a healthcare system. In addition to the use of CDS interventions, institutions should consider formal educational interventions targeted to clinicians with the most need to maximize appropriate albumin use.

## Data Availability

Aggregate data is presented in the manuscript. The IRB approval does not authorize sharing of study data with those outside of the research team.

## Declarations

### Funding

Dr. Umscheid’s contribution to this project was supported in part by the National Center for Research Resources, Grant UL1RR024134, which is now at the National Center for Advancing Translational Sciences, Grant UL1TR000003. The content of this paper is solely the responsibility of the authors and does not necessarily represent the official views of the NIH.

### Competing Interests

The authors report no potential financial conflicts of interest relevant to this article.

### Ethics Approval and Consent to Participate

The study received expedited approval and a Health Insurance Portability and Accountability Act (HIPAA) waiver from the University of Pennsylvania Institutional Review Board.

## Notes

### Competing Interest Statement

The authors have declared no competing interest.

